# NSAID use and clinical outcomes in COVID-19 patients: A 38-center retrospective cohort study

**DOI:** 10.1101/2021.04.13.21255438

**Authors:** Justin T. Reese, Ben Coleman, Lauren Chan, Hannah Blau, Tiffany J. Callahan, Luca Cappelletti, Tommaso Fontana, Katie Rebecca Bradwell, Nomi L. Harris, Elena Casiraghi, Giorgio Valentini, Guy Karlebach, Rachel Deer, Julie A. McMurry, Melissa A. Haendel, Christopher G. Chute, Emily Pfaff, Richard Moffitt, Heidi Spratt, Jasvinder Singh, Christopher J. Mungall, Andrew E. Williams, Peter N. Robinson

## Abstract

**Background:** Non-steroidal anti-inflammatory drugs (NSAIDs) are commonly used to reduce pain, fever, and inflammation but have been associated with complications in community-acquired pneumonia. Observations shortly after the start of the COVID-19 pandemic in 2020 suggested that ibuprofen was associated with an increased risk of adverse events in COVID-19 patients, but subsequent observational studies failed to demonstrate increased risk and in one case showed reduced risk associated with NSAID use.

**Methods:** A 38-center retrospective cohort study was performed that leveraged the harmonized, high-granularity electronic health record data of the National COVID Cohort Collaborative. A propensity-matched cohort of COVID-19 inpatients was constructed by matching cases (treated with NSAIDs) and controls (not treated) from 857,061 patients with COVID-19. The primary outcome of interest was COVID-19 severity in hospitalized patients, which was classified as: moderate, severe, or mortality/hospice. Secondary outcomes were acute kidney injury (AKI), extracorporeal membrane oxygenation (ECMO), invasive ventilation, and all-cause mortality at any time following COVID-19 diagnosis.

**Results:** Logistic regression showed that NSAID use was not associated with increased COVID-19 severity (OR: 0.57 95% CI: 0.53-0.61). Analysis of secondary outcomes using logistic regression showed that NSAID use was not associated with increased risk of all-cause mortality (OR 0.51 95% CI: 0.47-0.56), invasive ventilation (OR: 0.59 95% CI: 0.55-0.64), AKI (OR: 0.67 95% CI: 0.63-0.72), or ECMO (OR: 0.51 95% CI: 0.36-0.7). In contrast, the odds ratios indicate reduced risk of these outcomes, but our quantitative bias analysis showed E-values of between 1.9 and 3.3 for these associations, indicating that comparatively weak or moderate confounder associations could explain away the observed associations.

**Conclusions:** Study interpretation is limited by the observational design. Recording of NSAID use may have been incomplete. Our study demonstrates that NSAID use is not associated with increased COVID-19 severity, all-cause mortality, invasive ventilation, AKI, or ECMO in COVID-19 inpatients. A conservative interpretation in light of the quantitative bias analysis is that there is no evidence that NSAID use is associated with risk of increased severity or the other measured outcomes. Our findings are the largest EHR-based analysis of the effect of NSAIDs on outcome in COVID-19 patients to date. Our results confirm and extend analogous findings in previous observational studies using a large cohort of patients drawn from 38 centers in a nationally representative multicenter database.

## Background

As of November 2021, severe acute respiratory syndrome associated with coronavirus-2 (SARS-CoV-2) has infected more than 248 million people and caused more than 5 million deaths worldwide [1]. SARS-CoV-2 is the cause of the coronavirus disease of 2019 (COVID-19), a condition characterized by pneumonia, hyperinflammation, hypoxemic respiratory failure, a prothrombotic state, cardiac dysfunction, substantial mortality, and persistent morbidity in some survivors [2,3].

Non-steroidal anti-inflammatory drugs (NSAIDs) are a large and heterogeneous class of medications defined by their ability to inhibit cyclooxygenase (COX), an enzyme that catalyzes the conversion of arachidonic acid to prostaglandins. Due to their widespread use, NSAIDs are common causes of serious adverse events that frequently necessitate hospitalization [4]. NSAIDs have numerous potentially deleterious effects on immune function [5,6] and may also mask warning signs of severe infection such as fever and pain during the course of community-acquired pneumonia [7]. NSAID exposure in the early stage of community-acquired pneumonia has been associated with a delayed diagnosis and more severe clinical course,[8,9] but the quality of available research has been called into doubt and recent studies have failed to reproduce the proposed association [10,11]. In a mouse model of COVID-19, NSAID treatment reduced both the antibody and proinflammatory cytokine response to SARS-CoV-2 infection. However, the timing of NSAID treatment may be relevant. It is possible that early NSAID treatment may negatively impact the initiation of antiviral immune responses while later NSAID treatment could be beneficial by suppressing immune-driven pathology such as cytokine storm [12,13], but evidence for this supposition is lacking.

A study published early in the course of the pandemic suggested that ibuprofen use was associated with more severe COVID-19 outcomes [8,14]. However, several subsequent studies failed to demonstrate a significant association between NSAID use and adverse outcomes in COVID-19 patients [10,15–26]. A prospective, multicenter cohort study on 78,674 hospitalized COVID-19 patients across 255 health-care facilities in England, Scotland, and Wales showed that NSAID use was not associated with worse in-hospital mortality, critical care admission, requirement for invasive ventilation, requirement for non-invasive ventilation, requirement for oxygen, or occurrence of acute kidney injury (AKI) [27]. Finally, a study on OpenSAFELY, an English data analytics platform, showed no evidence of a difference in the risk of COVID-19-related death associated with current use of NSAIDs among 2,463,707 individuals, 536,423 of whom had recorded NSAID use. The same study demonstrated a lower risk of COVID-19-related death was associated with current use of NSAIDs in a cohort of 1,708,781 individuals with rheumatoid arthritis/osteoarthritis, 175,495 of whom had recorded NSAID use [28].

Theoretical considerations suggest a potentially deleterious effect of NSAIDs especially early in the clinical course of COVID-19. The widespread use of NSAIDs coupled with the difficulty of performing a randomized clinical trial on over-the-counter medications make it essential to assess the safety of this class of medication in different settings.

Here, we leverage data from the National COVID Cohort Collaborative (N3C), a centralized, harmonized, high-granularity electronic health record (EHR) repository to investigate potential associations of NSAID use in a large, multi-center database [29]. We investigated twelve NSAIDs (celecoxib, diclofenac, droxicam, etodolac, ketorolac, ibuprofen, indomethacin, lornoxicam, meloxicam, naproxen, piroxicam, tenoxicam) that were previously evaluated for their effect on COVID-19 severity in a smaller cohort earlier in the COVID-19 pandemic [27]. Our study focused on hospitalized patients with moderate and severe COVID-19. Our results showed significant associations of NSAID use with decreased all-cause mortality, COVID-19 severity, AKI, invasive ventilation, and extracorporeal membrane oxygenation (ECMO). Our analysis confirms and extends analogous findings from previous observational studies to a much larger cohort of patients drawn from 38 distinct centers in a nationally representative database.

## Methods

### Data analysis

Data analysis was performed using Palantir Foundry (Palantir Technologies Inc., Denver, Colorado). The analysis was structured as a directed acyclic graph of data transformations using the Foundry platform. Individual transformations were implemented using SQL, Python, or R code. Documentation of the source code is included in Supplementary Data.

### Design Overview, Settings, and Participants

Patient data were accessed through the N3C (covid.cd2h.org). N3C aggregates and harmonizes EHR data across 66 clinical organizations in the United States, including the Clinical and Translational Science Awards (CTSA) Program hubs. For this analysis, data were derived from the 38 centers that provided data for all predictors used in the regression analysis described below. Twenty-six centers did not provide data on Body Mass Index (BMI) and were not included in this study. N3C harmonizes data across four clinical data models and provides a unified analytical platform in which data are encoded using the Observational Medical Outcomes Partnership (OMOP) [30] version 5.3.1. N3C also provides shared phenotype definitions such as those for positive COVID-19 laboratory tests and COVID-19 clinical severity categories [29,31]. Our study analyzed only inpatients, as a preliminary analysis indicated that NSAID use among outpatients was likely to be incompletely captured (Supplementary Table S1).

For each of the twelve NSAIDs and the excluded medications (aspirin and acetaminophen), we constructed a codeset containing concept IDs representing all formulations of the medications using ATLAS (http://atlas-covid19.ohdsi.org/), the graphical user interface designed to construct cohorts and/or concept sets for the OMOP common data model [32]. Concept IDs for topical and ophthalmic NSAID preparations were excluded from these codesets. For each analyzed comorbidity, a codeset was constructed containing concept IDs representing the comorbidity in question. OMOP concept ID codes for all drugs and comorbidities used in this analysis are listed in Supplemental Tables S2 and S3.

Criteria for the current study were determined as follows. The COVID-19 positive cohort was defined as those patients with any encounter after January 1, 2020 and positive SARS-CoV-2 laboratory test (polymerase chain reaction or antigen). For this study, data from up to October 5, 2021 were included. COVID-19 positive patients whose drug era [33] for any of the 12 NSAIDs began on or before the initial date of COVID-19 diagnosis and continued for at least one day after COVID-19 diagnosis were included in the NSAID treated group. Patients with recorded use of aspirin and acetaminophen, which shares some modes of action with NSAIDs, were excluded from the analysis. As before, use of aspirin and acetaminophen was defined as a drug era for either of these drugs that began on or before the day of COVID-19 diagnosis and continued for at least one day. All other patients from the COVID-19 positive cohort were used as the control group in propensity matching. For each patient, comorbidities that were diagnosed before the day of diagnosis of COVID-19 were also recorded.

Only patients with complete records (no missing values for any covariate used in propensity matching or logistic regression) were included for further analysis. The most commonly missing data was BMI. Supplemental Figures S1-S2 show similar distributions of age and Charlson Comorbidity Index [34] in the presence or absence of BMI.

#### OUTCOMES

The primary outcome of interest was COVID-19 clinical severity of “severe” or “mortality/hospice”. Clinical severity was classified into three categories using the Clinical Progression Scale (CPS) established by the World Health Organization (WHO) for COVID-19 clinical research [35]: WHO severity 3); “moderate” (hospitalized without invasive ventilation, WHO severity 4-6); “severe” (hospitalized with invasive ventilation or ECMO, WHO severity 7-9); and “mortality/hospice” (hospital mortality or discharge to hospice, WHO Severity 10) [31]. In our study, severity grade 3 (moderate) was compared against severity grades 4 and 5 (severe or mortality/hospice). For the purposes of our study, the severity grades of mild and mild ED (emergency department) were not included. For the logistic regression analysis described below, patients were assigned to COVID-19 severity groups according to the maximum clinical severity during their index encounter [31], which was defined as the medical encounter during which a positive COVID-19 test was documented for the first time. Secondary outcomes were AKI, ECMO, invasive ventilation, and all-cause mortality at any time following COVID-19 diagnosis.

### Study design

**Figure 1.**
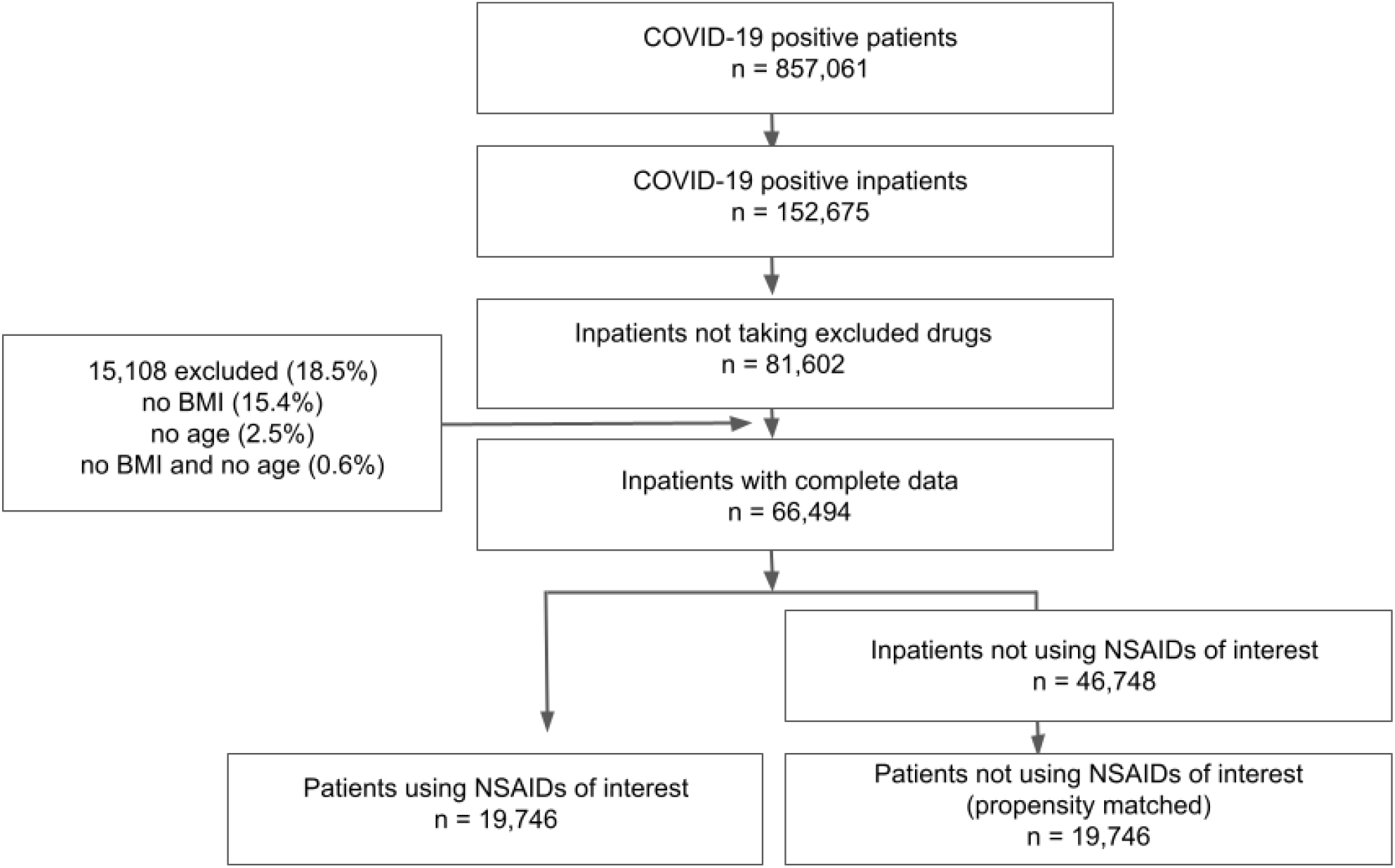
Definition of NSAID cohort and matched control cohort for analysis of the association of NSAID use with COVID-19 outcome.

#### Statistical analysis

We performed propensity matching using the “nearest” method implemented in the R *MatchIt* package (version 4.1.0). Each patient from the NSAID treated group was matched to the patient in the untreated group with the closest propensity score. The propensity formula included age, race, ethnicity, gender, smoking status, Charlson Comorbidity Index, and BMI, as well as the presence or absence of a diagnosis of the following comorbidities before COVID presentation: alcoholic liver damage, Alzheimer’s disease, cerebral infarction, chronic hepatitis, chronic respiratory disease, dementia associated with another disease, diabetes type 1, diabetes type 2, hepatic failure, hepatic fibrosis, hepatic steatosis, hypertension, hypertensive kidney disease, ischemic heart disease, lupus, malignant neoplasm (lymphoid hematopoietic related tissue), neoplasm, nicotine dependence, nonhypertensive chronic kidney disease, nonischemic heart disease, other liver disease, portal hypertension, psoriasis, rheumatoid arthritis, unspecified dementia, and vascular dementia.

To investigate the association of treatment and other covariates with COVID-19 severity, we performed logistic regression using the *glm* function in R. The dependent variable, COVID-19 severity, was coded as 0 for patients with “moderate” COVID-19 severity and 1 for patients with “severe”, or “mortality/hospice” COVID-19 severity [31]. We assessed the relationship between COVID-19 severity and NSAID use using logistic regression, using as additional predictors age, race, ethnicity, gender, smoking status, Charlson Comorbidity Index, and BMI, as well as the following comorbidities: alcoholic liver damage, Alzheimer’s disease, cerebral infarction, chronic respiratory disease, diabetes type 1, diabetes type 2, hepatic failure, hepatic fibrosis, hypertensive kidney disease, ischemic heart disease, lupus, malignant neoplasm (lymphoid hematopoietic related tissue), neoplasm, nicotine dependence, nonhypertensive chronic kidney disease, nonischemic heart disease, other liver disease, portal hypertension, psoriasis, unspecified dementia, and vascular dementia. For treatment with the medication, we recorded the estimate, the corresponding p-value, the odds ratio, and 95% confidence intervals.

We used the *EValue* R package (version 4.1.2) to determine the minimum strength of an unmeasured confounder in the logistic regression that would be required to change the conclusion that the treatment (NSAID use) was associated with the outcome in question. We treated the outcome of increased COVID-19 severity as a non-rare outcome (as it occurred more frequently than 15%), and death, invasive ventilation, AKI, and ECMO as rare outcomes (since the frequency of these outcomes was less than 15%) [36].

## Results

We evaluated 857,061 patients with COVID-19 in a retrospective study and evaluated twelve NSAIDs that were evaluated previously on a smaller cohort [27]. Individuals diagnosed with COVID-19 were then divided into those individuals treated with the medication (treated) and those who were not (controls). To reduce the effect of confounding, we performed propensity matching [37] to match NSAID-treated and control patients according to age, race, ethnicity, gender, smoking status, Charlson Comorbidity Index, BMI, and the presence of 24 comorbidities before COVID-19 presentation (Methods). Table 1 shows the composition of the cohort with respect to these covariates before and after propensity matching. After propensity matching, the standard mean difference between NSAID-treated and control groups for all covariates was less than 0.1.

**Table 1.** Characteristics of the COVID-19 positive cohort taking NSAIDs and the control cohort, before and after propensity matching. SMD: standardized mean difference between NSAID and control cohorts.

|  |  | Before Propensity Matching |  | After Propensity Matching |  |
| --- | --- | --- | --- | --- | --- |
|  | Treated | Control | SMD | Control | SMD |
| Age | 47.4 | 53.7 | -0.33 | 47.1 | 0.01 |
| Race |  |  |  |  |  |
| - Asian | 3.1% | 3.7% | -0.03 | 3.1% | 0.00 |
| - Black or African American | 21.6% | 20.9% | 0.02 | 22.3% | -0.02 |
| - Missing/Unknown | 25.1% | 23.3% | 0.04 | 24.8% | 0.01 |
| - Native Hawaiian or Other Pacific Islander | 0.2% | 0.3% | -0.01 | 0.2% | 0.00 |
| - Other | 0.9% | 0.9% | -0.01 | 0.8% | 0.01 |
| - White | 49.1% | 50.9% | -0.04 | 48.9% | 0.00 |
| Ethnicity |  |  |  |  |  |
| - Hispanic or Latino | 24.8% | 22.0% | 0.06 | 23.8% | 0.02 |
| - Missing/Unknown | 5.4% | 8.4% | -0.13 | 5.4% | 0.00 |
| - Not Hispanic or Latino | 69.8% | 69.6% | 0.00 | 70.8% | -0.02 |
| Gender |  |  |  |  |  |
| - Female | 59.7% | 50.1% | 0.20 | 59.1% | 0.01 |
| - Male | 40.3% | 49.8% | -0.20 | 40.9% | -0.01 |
| - Other | 0.0% | 0.0% | 0.01 | 0.0% | 0.00 |
| Smoking status |  |  |  |  |  |
| - Current or Former | 39.8% | 29.9% | 0.20 | 36.7% | 0.06 |
| - Non smoker | 60.2% | 70.1% | -0.20 | 63.3% | -0.06 |
| BMI | 3133.0% | 2974.2% | 0.18 | 3102.3% | 0.03 |
| Charlson Comorbidity Index | 98.1% | 143.5% | -0.23 | 96.3% | 0.01 |
| Alcoholic liver damage | 0.6% | 1.0% | -0.05 | 0.6% | 0.00 |
| Chronic hepatitis | 1.0% | 1.1% | -0.01 | 0.9% | 0.01 |
| Diabetes type 2 | 16.7% | 18.0% | -0.03 | 16.1% | 0.02 |
| Hepatic failure | 0.5% | 1.2% | -0.11 | 0.5% | 0.00 |
| Hypertension | 30.8% | 31.3% | -0.01 | 29.9% | 0.02 |
| Ischemic heart disease | 4.9% | 7.0% | -0.10 | 4.9% | 0.00 |
| Lupus | 0.7% | 0.6% | 0.02 | 0.7% | 0.00 |
| Malignant neoplasm (lymphoid hematopoietic related tissue) | 2.0% | 2.0% | 0.00 | 2.0% | 0.00 |
| Neoplasm | 19.4% | 17.2% | 0.06 | 19.2% | 0.00 |
| Nonischemic heart disease | 20.5% | 24.4% | -0.10 | 20.4% | 0.00 |
| Vascular dementia | 0.4% | 0.8% | -0.07 | 0.3% | 0.00 |
| Alzheimer's disease | 0.4% | 1.0% | -0.09 | 0.4% | 0.01 |
| Cerebral infarction | 1.7% | 2.9% | -0.10 | 1.6% | 0.01 |
| Chronic respiratory disease | 13.1% | 13.2% | 0.00 | 12.8% | 0.01 |
| Dementia associated with another disease | 1.0% | 1.5% | -0.05 | 0.9% | 0.01 |
| Diabetes type 1 | 1.4% | 1.5% | -0.01 | 1.4% | 0.00 |
| Hepatic fibrosis | 1.2% | 2.2% | -0.09 | 1.2% | 0.00 |
| Hepatic steatosis | 4.3% | 3.2% | 0.06 | 4.0% | 0.02 |
| Hypertensive kidney disease | 3.1% | 6.3% | -0.19 | 3.1% | 0.00 |
| Nicotine dependence | 9.8% | 8.9% | 0.03 | 9.6% | 0.01 |
| Nonhypertensive chronic kidney disease | 5.5% | 11.2% | -0.25 | 5.4% | 0.00 |
| Other liver disease | 5.2% | 5.4% | -0.01 | 4.8% | 0.02 |
| Portal hypertension | 0.5% | 1.2% | -0.10 | 0.5% | 0.00 |
| Rheumatoid arthritis | 1.9% | 1.3% | 0.04 | 1.8% | 0.01 |
| Unspecified dementia | 1.2% | 2.7% | -0.14 | 1.1% | 0.01 |
| Psoriasis | 1.0% | 0.8% | 0.01 | 0.9% | 0.00 |

The primary outcome was COVID-19 severity of moderate vs. severe or mortality/hospice (Table 2). No mild or mild ED cases were present in our treated or control groups. The incidence of moderate COVID-19 was higher among the NSAID cohort (91.3%) compared with the control cohort (86%), while the incidence of COVID-19 more severe than moderate (severe, or dead) was lower in the NSAID cohort (3.9% severe, 4.8% dead) compared to the control cohort (5.3% severe, 8.8% dead). Similarly, the incidence of invasive ventilation, AKI, and ECMO were all lower in the NSAID group compared to the control group.

**Table 2.** Outcomes in NSAID cohort and propensity-matched control cohort. Number of patients and percent of cohort for each outcome are shown. AKI: acute kidney injury; ECMO: extracorporeal membrane oxygenation.

|  | NSAID | Control |
| --- | --- | --- |
| COVID-19 severity |  |  |
| Moderate | 18023 (91.3%) | 16972 (86%) |
| Severe | 776 (3.9%) | 1047 (5.3%) |
| Dead | 947 (4.8%) | 1727 (8.8%) |
| Invasive ventilation | 1150 (5.8%) | 1867 (9.5%) |
| AKI | 1729 (8.8%) | 2437 (12.3%) |
| ECMO | 54 (0.27%) | 109 (0.55%) |

We observed a significant association between NSAID use and lower COVID-19 severity when controlling for age, race, ethnicity, gender, smoking status, Charlson comorbidity, BMI, and 22 other comorbidities (OR 0.57, Table 3).

**Table 3.** Association of NSAID use with COVID-19 outcomes as measured by logistic regression. AKI: acute kidney injury; ECMO: extracorporeal membrane oxygenation.

|  | OR (95% CI) | p value |
| --- | --- | --- |
| COVID severity (severe or dead) | 0.57 (0.53-0.61) | < 0.0001 |
| Mortality/hospice | 0.51 (0.47-0.56) | < 0.0001 |
| Invasive ventilation | 0.59 (0.55-0.64) | < 0.0001 |
| AKI | 0.67 (0.63-0.72) | < 0.0001 |
| ECMO | 0.51 (0.36-0.7) | < 0.0001 |

We further investigated the association of NSAID use with four secondary outcomes: AKI, ECMO, invasive ventilation, and all-cause mortality at any time following COVID-19 diagnosis. The characteristics of the NSAID treated and control cohorts with respect to outcomes are shown in Table 2. We analyzed the association of NSAID use with these four other secondary outcomes using logistic regression (Table 3). NSAID use was significantly associated with fewer incidents of death (OR 0.51), invasive ventilation (OR 0.59), AKI (OR 0.67), and ECMO (OR 0.51).

### Quantitative bias analysis

We calculated the E-value for the observed values of the odds ratio to assess the sensitivity of our findings to uncorrected confounders [36]. To fully explain the association of NSAID use with decreased COVID-19 severity, an unmeasured confounder would need to be associated with both the treatment and the outcome with an odds ratio of at least 1.88 above and beyond the confounders included in the regression. Likewise, to fully explain the association of NSAID use with fewer adverse secondary outcomes, a confounder would need to be associated with both the treatment and the outcome with an odds ratio of at least 3.3 (death), 2.8 (invasive ventilation), 2.3 (AKI), and 3.3 (ECMO).

## Discussion

To our knowledge, randomized clinical trial data investigating potential beneficial or deleterious effects of NSAIDs on the course of COVID-19 are not available. Previous observational studies have failed to show an association of exposure to NSAIDS with risk of hospital admission, severe clinical course, or death in COVID-19 patients [38]. Our study is the second-largest to be performed to date and the largest to be performed as a multi-center American study (Supplemental Table S4). The largest study leveraged data in the OpenSAFELY platform, which includes information from primary care practices in England, including pseudonymised data such as coded diagnoses, prescribed medications, and physiological parameters. Our study represents the largest multi-center study with data harmonized from multiple EHR data sources. Our findings did not show an association in hospitalized COVID-19 patients between NSAID use and increased COVID-19 severity, or increased risk of invasive ventilation, AKI, ECMO, and all-cause mortality. In contrast, we identified a significant association between NSAID use and *decreased* risk of these outcomes. These results are in accordance with those of some previous studies: NSAID use was reported less frequently among hospitalized patients than non-hospitalized patients [16]; a study on 1305 hospitalized COVID-19 patients showed that use of NSAIDs prior to hospitalization was associated with lower odds of mortality as assessed by multivariate regression analysis [25]; finally, the OpenSAFELY study demonstrated a lower risk of COVID-19-related death was associated with current use of NSAIDs in individuals with rheumatoid arthritis/osteoarthritis [28]. However, we observed E-values for these associations in the range of 1.9-3.3, indicating that comparatively weak or moderate confounder associations could explain away the observed associations. We interpret the results of our study as providing additional evidence for the lack of a detrimental effect of NSAID use prior to hospital admission on the severity and other deleterious outcomes of COVID-19. Future work will be required to investigate a potential beneficial effect.

### Strengths and limitations of this study

Our dataset is derived from over 38 institutions across the country with 857,061 cases of COVID-19, and thus is a representative sample of the COVID-19 positive population in the United States. Observational studies such as retrospective EHR cohort analyses are subject to confounding. In the case of our study, the decision of whether to treat a patient with an NSAID could in principle be correlated with the outcome of interest (COVID-19 severity). We applied propensity matching to mitigate confounding, and used E-values to measure the strength of a confounder that would be required to change the conclusion of our analysis. However, in observational studies a risk of residual confounding persists because the efficacy of propensity matching is limited to known and measured factors. Exposure to the drugs of interest, most of which are available without a prescription, was likely to be captured incompletely in the EHR data used in the analysis. Thus, it is possible that there was unrecorded use of NSAIDs in the untreated group. Our study analyzed only inpatients, whose NSAID use is likely more to be completely captured by EHR data.

## Conclusions

The results of our observational study failed to demonstrate a significant association of the use of twelve NSAIDs with increased clinical severity, invasive ventilation, and AKI. Our study provides additional support to the notion that NSAIDs are safe for use in patients with COVID-19.

## Data Availability

Data are available through the N3C Data Enclave.

https://ncats.nih.gov/n3c/resources

## Declarations

### Ethics approval and consent to participate

The N3C data transfer to NCATS is performed under a Johns Hopkins University Reliance Protocol #IRB00249128 or individual site agreements with NIH. The N3C Data Enclave is managed under the authority of the NIH; information can be found at https://ncats.nih.gov/n3c/resources.

### Consent for publication

Not applicable.

### Availability of data and materials

Source code used in this study is available in the supplemental data. Data are available within the N3C Enclave.

### Authors’ contributions

Contributions are organized according to contribution roles as follows:

-data curation: Katie Rebecca Bradwell, Lauren Chan, Christopher G. Chute
-data integration: Justin T. Reese, Christopher G. Chute, Christopher J. Mungall
-data quality assurance: Tiffany J. Callahan, Lauren Chan
-data visualization: Justin T. Reese, Lauren Chan, Julie A. McMurry
-clinical subject matter expertise: Tiffany J. Callahan, Ben Coleman, Rachel Deer, Jasvinder Singh, Peter N. Robinson
-manuscript drafting: Justin T. Reese, Hannah Blau, Lauren Chan, Melissa A. Haendel, Jasvinder Singh, Heidi Spratt, Peter N. Robinson
-project management: Justin T. Reese, Nomi L. Harris, Christopher G. Chute
-clinical data model expertise: Justin T. Reese, Katie Rebecca Bradwell, Melissa A. Haendel, Christopher G. Chute, Andrew E. Williams
-funding acquisition: Melissa A. Haendel, Christopher G. Chute
-N3C Phenotype definition: Christopher G. Chute, Emily Pfaff, Richard Moffitt
-biological subject matter expertise: Justin T. Reese, Melissa A. Haendel
-statistical analysis: Justin T. Reese, Heidi Spratt, Andrew E. Williams, Peter N. Robinson
-governance: Christopher G. Chute
-regulatory oversight / admin: Christopher G. Chute
-project evaluation: Justin T. Reese, Christopher G. Chute
-critical revision of the manuscript for important intellectual content: Justin T. Reese, Katie Rebecca Bradwell, Tiffany J. Callahan, Nomi L. Harris, Rachel Deer, Julie A. McMurry, Christopher G. Chute, Jasvinder Singh, Heidi Spratt, Andrew E. Williams, Peter Robinson, Richard Moffitt, Emily Pfaff

## Acknowledgements

The analyses described in this publication were conducted with data or tools accessed through the NCATS N3C Data Enclave (covid.cd2h.org/enclave) and supported by NCATS U24 TR002306. This research was possible because of the patients whose information is included within the data from participating organizations:

https://ncats.nih.gov/n3c/resources/data-contribution/data-transfer-agreement-signatories and scientists who have contributed to the ongoing development of this community resource (doi.org/10.5281/zenodo.3979622). Authorship was determined using ICMJE recommendations.

We are grateful to the following data partners for providing data, made possible by supporting grants:

Stony Brook University — U24TR002306 • University of Oklahoma Health Sciences Center — U54GM104938: Oklahoma Clinical and Translational Science Institute (OCTSI) • West Virginia University — U54GM104942: West Virginia Clinical and Translational Science Institute (WVCTSI) • University of Mississippi Medical Center — U54GM115428: Mississippi Center for Clinical and Translational Research (CCTR) • University of Nebraska Medical Center — U54GM115458: Great Plains IDeA-Clinical & Translational Research • Maine Medical Center — U54GM115516: Northern New England Clinical & Translational Research (NNE-CTR) Network • Wake Forest University Health Sciences — UL1TR001420: Wake Forest Clinical and Translational Science Institute • Northwestern University at Chicago — UL1TR001422: Northwestern University Clinical and Translational Science Institute (NUCATS) • University of Cincinnati — UL1TR001425: Center for Clinical and Translational Science and Training • The University of Texas Medical Branch at Galveston — UL1TR001439: The Institute for Translational Sciences • Medical University of South Carolina — UL1TR001450: South Carolina Clinical & Translational Research Institute (SCTR) • University of Massachusetts Medical School Worcester — UL1TR001453: The UMass Center for Clinical and Translational Science (UMCCTS) • University of Southern California — UL1TR001855: The Southern California Clinical and Translational Science Institute (SC CTSI) • Columbia University Irving Medical Center — UL1TR001873: Irving Institute for Clinical and Translational Research • George Washington Children’s Research Institute — UL1TR001876: Clinical and Translational Science Institute at Children’s National (CTSA-CN) • University of Kentucky — UL1TR001998: UK Center for Clinical and Translational Science • University of Rochester — UL1TR002001: UR Clinical & Translational Science Institute • University of Illinois at Chicago — UL1TR002003: UIC Center for Clinical and Translational Science • Penn State Health Milton S. Hershey Medical Center — UL1TR002014: Penn State Clinical and Translational Science Institute • The University of Michigan at Ann Arbor — UL1TR002240: Michigan Institute for Clinical and Health Research • Vanderbilt University Medical Center — UL1TR002243: Vanderbilt Institute for Clinical and Translational Research • University of Washington — UL1TR002319: Institute of Translational Health Sciences • Washington University in St. Louis — UL1TR002345: Institute of Clinical and Translational Sciences • Oregon Health & Science University — UL1TR002369: Oregon Clinical and Translational Research Institute • University of Wisconsin-Madison — UL1TR002373: UW Institute for Clinical and Translational Research • Rush University Medical Center — UL1TR002389: The Institute for Translational Medicine (ITM) • The University of Chicago — UL1TR002389: The Institute for Translational Medicine (ITM) • University of North Carolina at Chapel Hill — UL1TR002489: North Carolina Translational and Clinical Science Institute • University of Minnesota — UL1TR002494: Clinical and Translational Science Institute • Children’s Hospital Colorado — UL1TR002535: Colorado Clinical and Translational Sciences Institute • The University of Iowa — UL1TR002537: Institute for Clinical and Translational Science • The University of Utah — UL1TR002538: Uhealth Center for Clinical and Translational Science • Tufts Medical Center — UL1TR002544: Tufts Clinical and Translational Science Institute • Duke University — UL1TR002553: Duke Clinical and Translational Science Institute • Virginia Commonwealth University — UL1TR002649: C. Kenneth and Dianne Wright Center for Clinical and Translational Research • The Ohio State University — UL1TR002733: Center for Clinical and Translational Science • The University of Miami Leonard M. Miller School of Medicine — UL1TR002736: University of Miami Clinical and Translational Science Institute • University of Virginia — UL1TR003015: iTHRIV Integrated Translational health Research Institute of Virginia • Carilion Clinic — UL1TR003015: iTHRIV Integrated Translational health Research Institute of Virginia • University of Alabama at Birmingham — UL1TR003096: Center for Clinical and Translational Science • Johns Hopkins University — UL1TR003098: Johns Hopkins Institute for Clinical and Translational Research • University of Arkansas for Medical Sciences — UL1TR003107: UAMS Translational Research Institute • Nemours — U54GM104941: Delaware CTR ACCEL Program • University Medical Center New Orleans — U54GM104940: Louisiana Clinical and Translational Science (LA CaTS) Center • University of Colorado Denver, Anschutz Medical Campus — UL1TR002535: Colorado Clinical and Translational Sciences Institute • Mayo Clinic Rochester — UL1TR002377: Mayo Clinic Center for Clinical and Translational Science (CCaTS) • Tulane University — UL1TR003096: Center for Clinical and Translational Science • Loyola University Medical Center — UL1TR002389: The Institute for Translational Medicine (ITM) • Advocate Health Care Network — UL1TR002389: The Institute for Translational Medicine (ITM) • OCHIN — INV-018455: Bill and Melinda Gates Foundation grant to Sage Bionetworks

## References

1. COVID-19 Map - Johns Hopkins Coronavirus Resource Center [Internet]. [cited 2021 Nov 15]. Available from: https://coronavirus.jhu.edu/map.html

2. Ortiz-Prado E, Simbaña-Rivera K, Gómez-Barreno L, Rubio-Neira M, Guaman LP, Kyriakidis NC, et al. Clinical, molecular, and epidemiological characterization of the SARS-CoV-2 virus and the Coronavirus Disease 2019 (COVID-19), a comprehensive literature review. Diagn Microbiol Infect Dis. 2020;98:115094.

3. Deer RR, Rock MA, Vasilevsky N, Carmody L, Rando H, Anzalone AJ, et al. Characterizing Long COVID: Deep Phenotype of a Complex Condition. EBioMedicine. 2021;74:103722.

4. Pirmohamed M, James S, Meakin S, Green C, Scott AK, Walley TJ, et al. Adverse drug reactions as cause of admission to hospital: prospective analysis of 18 820 patients. BMJ. 2004;329:15–9.

5. Graham NM, Burrell CJ, Douglas RM, Debelle P, Davies L. Adverse effects of aspirin, acetaminophen, and ibuprofen on immune function, viral shedding, and clinical status in rhinovirus-infected volunteers. J Infect Dis. 1990;162:1277–82.

6. Bancos S, Bernard MP, Topham DJ, Phipps RP. Ibuprofen and other widely used non-steroidal anti-inflammatory drugs inhibit antibody production in human cells. Cell Immunol. 2009;258:18–28.

7. Torjesen I. Ibuprofen can mask symptoms of infection and might worsen outcomes, says European drugs agency. BMJ. 2020;369:m1614.

8. Micallef J, Soeiro T, Jonville-Béra A-P, French Society of Pharmacology, Therapeutics (SFPT). Non-steroidal anti-inflammatory drugs, pharmacology, and COVID-19 infection. Therapie. 2020;75:355–62.

9. Voiriot G, Dury S, Parrot A, Mayaud C, Fartoukh M. Nonsteroidal antiinflammatory drugs may affect the presentation and course of community-acquired pneumonia. Chest. 2011;139:387–94.

10. Vaja R, Chan JSK, Ferreira P, Harky A, Rogers LJ, Gashaw HH, et al. The COVID-19 ibuprofen controversy: A systematic review of NSAIDs in adult acute lower respiratory tract infections. Br J Clin Pharmacol. 2021;87:776–84.

11. Lund LC, Reilev M, Hallas J, Kristensen KB, Thomsen RW, Christiansen CF, et al. Association of Nonsteroidal Anti-inflammatory Drug Use and Adverse Outcomes Among Patients Hospitalized With Influenza. JAMA Netw Open. 2020;3:e2013880.

12. Chen Jennifer S., Alfajaro Mia Madel, Chow Ryan D., Wei Jin, Filler Renata B., Eisenbarth Stephanie C., et al. Nonsteroidal Anti-inflammatory Drugs Dampen the Cytokine and Antibody Response to SARS-CoV-2 Infection. J Virol. American Society for Microbiology; 95:e00014–21.

13. Fajgenbaum DC, June CH. Cytokine Storm. N Engl J Med. 2020;383:2255–73.

14. Day M. Covid-19: ibuprofen should not be used for managing symptoms, say doctors and scientists. BMJ. 2020;368:m1086.

15. Choi MH, Ahn H, Ryu HS, Kim B-J, Jang J, Jung M, et al. Clinical Characteristics and Disease Progression in Early-Stage COVID-19 Patients in South Korea. J Clin Med Res [Internet]. 2020;9. Available from: http://dx.doi.org/10.3390/jcm9061959

16. Gianfrancesco M, Hyrich KL, Al-Adely S, Carmona L, Danila MI, Gossec L, et al. Characteristics associated with hospitalisation for COVID-19 in people with rheumatic disease: data from the COVID-19 Global Rheumatology Alliance physician-reported registry. Ann Rheum Dis. 2020;79:859–66.

17. Rinott E, Kozer E, Shapira Y, Bar-Haim A, Youngster I. Ibuprofen use and clinical outcomes in COVID-19 patients. Clin Microbiol Infect. 2020;26:1259.e5–1259.e7.

18. Bruce E, Barlow-Pay F, Short R, Vilches-Moraga A, Price A, McGovern A, et al. Prior Routine Use of Non-Steroidal Anti-Inflammatory Drugs (NSAIDs) and Important Outcomes in Hospitalised Patients with COVID-19. J Clin Med Res [Internet]. 2020;9. Available from: http://dx.doi.org/10.3390/jcm9082586

19. Lund LC, Kristensen KB, Reilev M, Christensen S, Thomsen RW, Christiansen CF, et al. Adverse outcomes and mortality in users of non-steroidal anti-inflammatory drugs who tested positive for SARS-CoV-2: A Danish nationwide cohort study. PLoS Med. 2020;17:e1003308.

20. Kragholm K, Gerds TA, Fosbøl E, Andersen MP, Phelps M, Butt JH, et al. Association Between Prescribed Ibuprofen and Severe COVID-19 Infection: A Nationwide Register-Based Cohort Study. Clin Transl Sci. 2020;13:1103–7.

21. Abu Esba LC, Alqahtani RA, Thomas A, Shamas N, Alswaidan L, Mardawi G. Ibuprofen and NSAID Use in COVID-19 Infected Patients Is Not Associated with Worse Outcomes: A Prospective Cohort Study. Infect Dis Ther. 2021;10:253–68.

22. Alamdari NM, Afaghi S, Rahimi FS, Tarki FE, Tavana S, Zali A, et al. Mortality Risk Factors among Hospitalized COVID-19 Patients in a Major Referral Center in Iran. Tohoku J Exp Med. 2020;252:73–84.

23. Chandan JS, Zemedikun DT, Thayakaran R, Byne N, Dhalla S, Acosta-Mena D, et al. Nonsteroidal Antiinflammatory Drugs and Susceptibility to COVID-19. Arthritis Rheumatol. 2021;73:731–9.

24. Hwang J-M, Kim J-H, Park J-S, Chang MC, Park D. Neurological diseases as mortality predictive factors for patients with COVID-19: a retrospective cohort study. Neurol Sci. 2020;41:2317–24.

25. Imam Z, Odish F, Gill I, O’Connor D, Armstrong J, Vanood A, et al. Older age and comorbidity are independent mortality predictors in a large cohort of 1305 COVID-19 patients in Michigan, United States. J Intern Med. 2020;288:469–76.

26. Sahai A, Bhandari R, Godwin M, McIntyre T, Chung MK, Iskandar J-P, et al. Effect of aspirin on short-term outcomes in hospitalized patients with COVID-19. Vasc Med. 2021;26:626–32.

27. Drake TM, Fairfield CJ, Pius R, Knight SR, Norman L, Girvan M, et al. Non-steroidal anti-inflammatory drug use and outcomes of COVID-19 in the ISARIC Clinical Characterisation Protocol UK cohort: a matched, prospective cohort study. Lancet Rheumatol. 2021;3:e498–506.

28. Wong AY, MacKenna B, Morton CE, Schultze A, Walker AJ, Bhaskaran K, et al. Use of non-steroidal anti-inflammatory drugs and risk of death from COVID-19: an OpenSAFELY cohort analysis based on two cohorts. Ann Rheum Dis [Internet]. 2021; Available from: http://dx.doi.org/10.1136/annrheumdis-2020-219517

29. Haendel MA, Chute CG, Bennett TD, Eichmann DA, Guinney J, Kibbe WA, et al. The National COVID Cohort Collaborative (N3C): Rationale, design, infrastructure, and deployment. J Am Med Inform Assoc. 2021;28:427–43.

30. Voss EA, Makadia R, Matcho A, Ma Q, Knoll C, Schuemie M, et al. Feasibility and utility of applications of the common data model to multiple, disparate observational health databases. J Am Med Inform Assoc. 2015;22:553–64.

31. Bennett TD, Moffitt RA, Hajagos JG, Amor B, Anand A, Bissell MM, et al. The National COVID Cohort Collaborative: Clinical Characterization and Early Severity Prediction. medRxiv [Internet]. 2021; Available from: http://dx.doi.org/10.1101/2021.01.12.21249511

32. Maier C, Kapsner LA, Mate S, Prokosch H-U, Kraus S. Patient Cohort Identification on Time Series Data Using the OMOP Common Data Model. Appl Clin Inform. 2021;12:57–64.

33. Hripcsak G, Duke JD, Shah NH, Reich CG, Huser V, Schuemie MJ, et al. Observational Health Data Sciences and Informatics (OHDSI): Opportunities for Observational Researchers. Stud Health Technol Inform. 2015;216:574–8.

34. Charlson ME, Pompei P, Ales KL, MacKenzie CR. A new method of classifying prognostic comorbidity in longitudinal studies: development and validation. J Chronic Dis. 1987;40:373–83.

35. WHO Working Group on the Clinical Characterisation and Management of COVID-19 infection. A minimal common outcome measure set for COVID-19 clinical research. Lancet Infect Dis. 2020;20:e192–7.

36. VanderWeele TJ, Ding P. Sensitivity Analysis in Observational Research: Introducing the E-Value. Ann Intern Med. 2017;167:268–74.

37. Zhang Z. Propensity score method: a non-parametric technique to reduce model dependence. Ann Transl Med. 2017;5:7.

38. Moore N, Bosco-Levy P, Thurin N, Blin P, Droz-Perroteau C. NSAIDs and COVID-19: A Systematic Review and Meta-analysis. Drug Saf. 2021;44:929–38.

