## Supplemental Data for "NSAID use and clinical outcomes in COVID-19 patients: A 38-center retrospective cohort study"

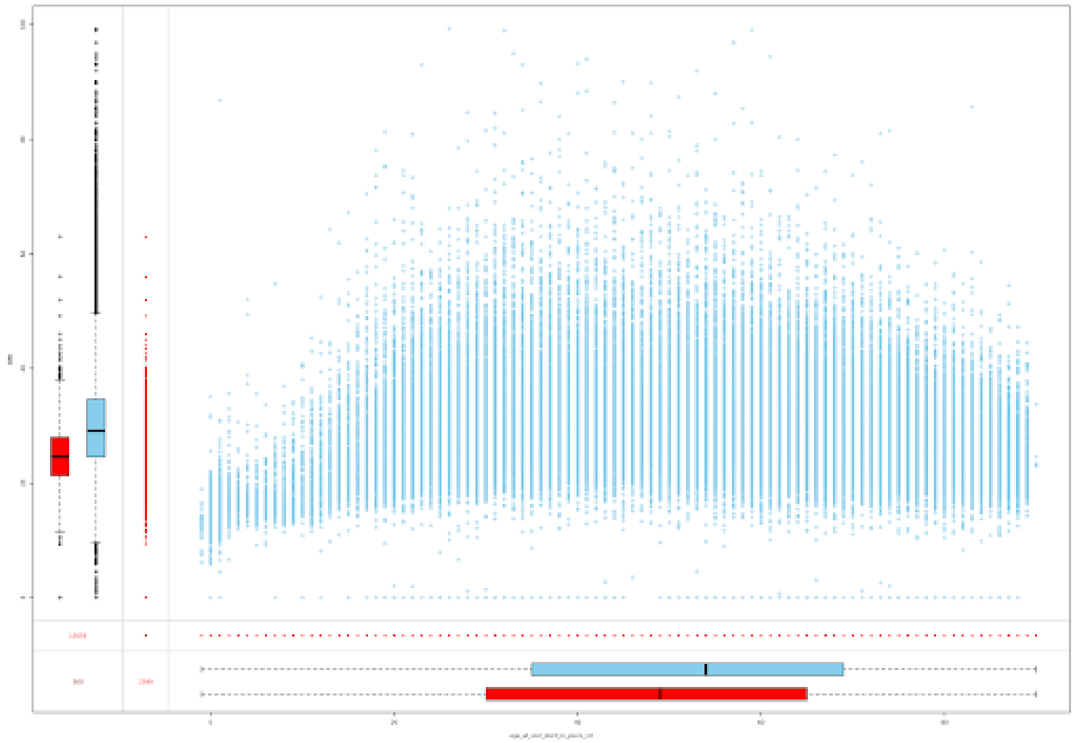

**Figure S1. Missingness analysis of BMI with respect to age**. The marginplot method in the VIM R package was used to analyze the distribution of BMI for patients with missing or non-missing age data, and the distribution of age for patients with missing or non-missing BMI. A box plot summarizing BMI in patients for whom age data is missing (red) or present is shown on the y-axis. A box plot summarizing age of patients for whom BMI is missing (red) or present (blue) is shown on the x-axis. The distribution of age data for patients with missing BMI was similar to that for patients with non-missing BMI. Likewise, the distribution of BMI data for patients with missing age was similar to that for patients with non-missing age. The plot suggests BMI and age data are missing completely at random.

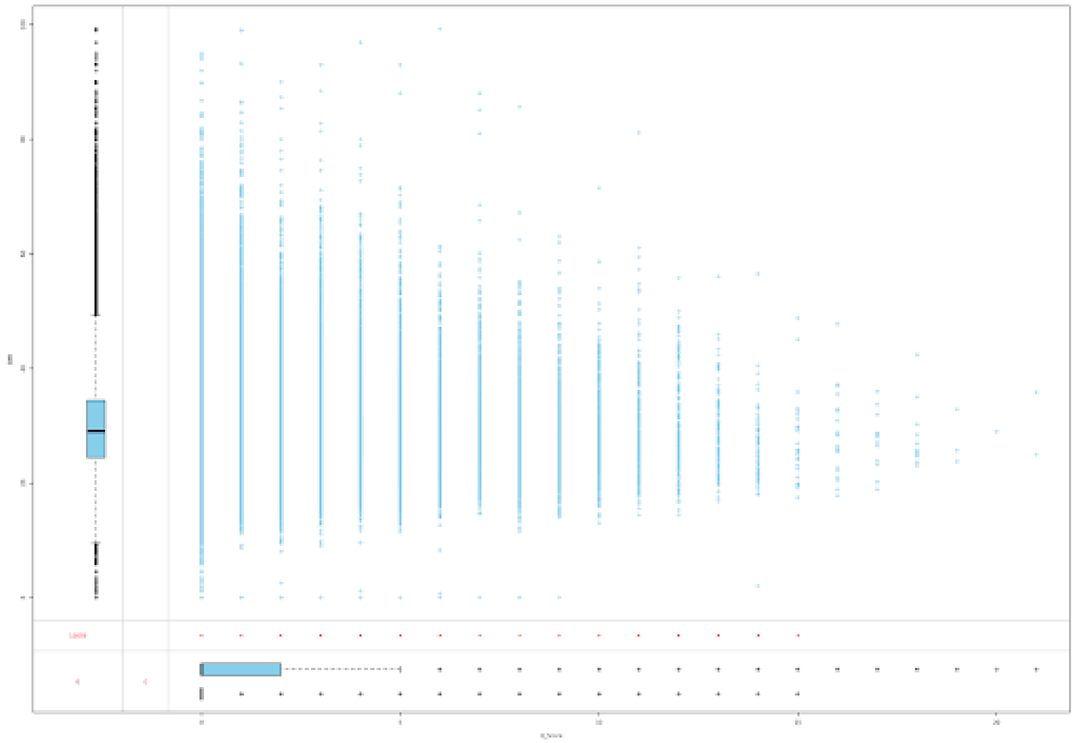

**Figure S2. Missingness analysis of BMI with respect to Charleson severity index score**. See Supplemental Figure S1 for explanations.

| COVID-19 severity | Percent of patients taking NSAIDs |
|---|---|
| Mild | 3.6% |
| Mild ED | 16.6% |
| Moderate | 16.2% |
| Severe | 9.4% |
| Mortality/hospice | 5.9% |

**Table S1. NSAID use by COVID-19 severity among the unmatched COVID-19 positive cohort.** NSAID use is likely incompletely captured disproportionately for outpatients and patients with less severe COVID-19, which may cause residual confounding.

| Medication | Concept id |
|---|---|
| ibuprofen | 1177480 |
| ketorolac | 1136980 |
| diclofenac | 1124300 |
| celecoxib | 1118084 |
| indomethacin | 1178663 |
| naproxen | 1115008 |
| etodolac | 1195492 |
| lornoxicam | 19049709 |
| tenoxicam | 19041220 |
| piroxicam | 1146810 |
| droxicam | 19056645 |
| meloxicam | 1150345 |
| aspirin | 1112807 |
| acetaminophen | 1125315 |

**Table S2. OMOP concept IDs for medications.** For each medication, the concept ID shown and all its descendants, excluding concepts representing topical and ophthalmic preparations, were used for analysis.

| Condition | Concept id |
|---|---|
| Alcoholic liver damage | 201612 |
| Chronic hepatitis | 200763 (excluded 4245975 and 201612) |
| Diabetes type 2 | 201826 |
| Hepatic failure | 4245975 (excluded 201612) |
| Hypertension | 316866 |
| Ischemic heart disease | 4185932 |
| Lupus | 255891 |
| Malignant neoplasm (lymphoid hematopoietic related tissue) | 4147164 |
| Neoplasm | 438112 (excluded 4147164) |
| Nonischemic heart disease | 321588 (excluded 4185932) |
| Vascular dementia | 443605 |
| Alzheimer's disease | 378419 (excluded 443605) |
| Cerebral infarction | 443454 |
| Chronic respiratory disease | 4063381 |
| Dementia associated with another disease | 374888 (excluded 378419 and 443605) |
| Diabetes type 1 | 201254 |
| Hepatic fibrosis | 4267417 (excluded 201612, 4245975, and 200763) |
| Hepatic steatosis | 4059290 (excluded 201612, 4245975, 200763, and 4267417) |
| Hypertensive kidney disease | 44782429 |
| Nicotine dependence | 4209423 |
| Nonhypertensive chronic kidney disease | 46271022 (excluded 44782429) |
| Other liver disease | 194984 (excluded 201612, 4245975, 200763, 4267417, 4059290, 200451, and 192680) |
| Portal hypertension | 192680 |
| Rheumatoid arthritis | 80809 (excluded 36684997) |
| Unspecified dementia | 4182210 (excluded 443605, 378419, and 374888) |
| Psoriasis | 140168 |

**Table S3. OMOP concept IDs for conditions.** For each condition, the concept ID shown and all its descendants were used for analysis.

| Study | Cohort (n) | On NSAIDs | Main findings |
|---|---|---|---|
| Abu Esba 2021 [1] | 503 | 146 | No association between ibuprofen or any other NSAID and worse COVID-19 outcomes |
| Alamdari 2020 [2] | 459 | 37 | NSAID use had no substantial impact on mortality. |
| Bruce 2020 [3] | 1222 | 54 | No association between prior NSAID use and time to mortality or length of stay. |
| Chandan2021 [4] | 25,659 | 13,202 | No increase in mortality among patients with osteoarthritis in a primary care setting. |
| Choi 2020 [5] | 293 | 8 | Ibuprofen was not a risk factor associated with disease progression. |
| Drake 2021 [6] | 78,674 | 4211 | NSAID use was not associated with worse in-hospital mortality, critical care admission, requirement for invasive ventilation, requirement for non-invasive ventilation, requirement for oxygen, or occurrence of acute kidney injury. |
| Gianfrancesco 2020 [7] | 531 | 111 | NSAID use not associated with hospitalisation status. |
| Gupta 2020 [8] | 2215 | 99 | No significant association with 28-day mortality |
| Hwang 2020 [9] | 103 | 5 | NSAID use showed no statistically significant difference in death rate. |
| Imam 2020 [10] | 1305 | 466 | Patients using NSAIDs prior to hospitalization hadlower odds of mortality. |
| Kragholm 2020 [11] | 4002 | 264 | No significant association between recent ibuprofen prescription claims and severe trajectory of COVID-19. |
| Lund 2020 [12] | 9236 | 248 | NSAIDs not associated with 30-day mortality, hospitalization, ICU admission, mechanical ventilation, or renal replacement therapy. |
| Park 2021 [13] | 7590 | 398 | NSAID use not associated with mortality or ventilator care in Covid-19 patients. |
| Rinott 2020 [14] | 403 | 87 | Ibuprofen use not associated with worse clinical outcomes, compared with paracetamol or no antipyretic. |
| Sahai 2021 [15] | 911 | 465 | NSAIDs use showed no significant differences in clinical covariates |
| Wong 2021 (#1) [16] | 2,463,707 | 536,423 | No evidence of difference in risk of COVID-19 related death associated with current use of NSAIDs |
| Wong 2021 (#2) [16] | 1,708,781 | 175,495 | In cohort of people with rheumatoid arthritis/osteoarthritis, a lower risk of COVID-19 related death was associated with current use of NSAID versus non-use. |

**Table S4.** Previously published studies on potential asosciations of NSAID use with outcome in COVID-19.

### S1 Supplemental Note 1

This project was conducted in the National Institute of Health (NIH) N3C Data Enclave on the Palantir Foundry platform (Palantir Technologies Inc., Denver, Colorado). This platform organizes code in nodes (transformations) that form a directed acyclic graph and cannot easily be presented in linearized fashion. The purpose of this documentation is to illustrate the approach taken by our analysis and to promote reproducibility and extensibility on the Palantir platform [17, 18] or (with appropriate changes) on other platforms.

The Palantir platform enables each node to use a different programming language. We chose SQL, Python, and R according to which technique was best adapted to the task at hand. Spark SQL (version 3.0.2-palantir.24), Python version 3.6.10, and R version 3.5.1 (2018-07-02) were used.

The N3C Data Enclave (hereafter "Enclave") contains data from over 2.4 million COVID-19 positive patients from 64 health systems in the United States. The dataset utilized in this study was frozen on October 5, 2021. Data were harmonized, integrated, and mapped with the Observational Medical Outcomes Partnership (OMOP) 5.3.1 vocabulary.

For clarity, we have omitted most of the code that was used to check and output results and concentrate on the code that performed the analysis.

#### S1.1 Inputs

The inputs for our analysis included OMOP tables as well as some tables provided by the Palantir platform with processed data.

##### S1.1.1 OMOP Tables

The following OMOP tables were used for our analysis (Table S5). We refer the reader to the original OMOP documentation for more information details [19].

| Table | Summary |
|---|---|
| `drug_era` | Represents a span of time when a patient is assumed to be exposed to a particular drug (active ingredient). |
| `condition_occurrence` | Dates when a condition is considered to have started and (if applicable) ended |

**Table S5.** OMOP tables used in this study.

##### S1.1.2 Tables offered by the Enclave

The following Enclave-specific tables were used for our analysis.

##### S1.1.3 concept set members

This table defines the relations between the codesets and concept sets (Table S6).

| codeset_id | concept_id | concept_set_name | concept_name |
|---|---|---|---|
| 837398380 | 436540 | Fracture | Open fracture axis |
| 837398380 | 436824 | Fracture | Open fracture of cervical vertebra without spinal cord injury |
| 837398380 | 436826 | Fracture | Closed fracture of shaft of radius |
| 837398380 | 436832 | Fracture | Closed fracture of sternum |
| 837398380 | 437117 | Fracture | Open fracture of intracapsular section of femur |
| (...) | (...) | (...) | (...) |

**Table S6. concept_set_members table**. Some examples from this table, which is used to coordinate codesets created by N3C Enclave members (often using the OMOP/OHDSI Atlas tool). The table has three addition columns (not shown here) that specify the version and were used to extract the latest versions for analysis.

#### S1.2 complete patient table with derived scores

This table is prepared as described [18], and contains derived information that was used to provide information about some of the covariates used in our analysis (Table S7).

| Column | data type | comment |
|---|---|---|
| person_id | string | unique patient identifier |
| data_partner_id | integer | identifier of N3C contributing center |
| visit_concept_id | integer | e.g., 9201 (Inpatient Visit) |
| visit_start_date | Date | The date of the initial medical encounter for COVID-19 or control condition |
| visit_concept_name | string | concept name corresponding to visit_concept_id |
| visit_occurrence_id | string | identifier for a visit |
| AKI_in_hospital | string | "Yes" or null |
| ECMO | string | "ECMO" or null |
| Invasive_Ventilation | string | "Invasive Ventilation" or null |
| positive_covid_test | string | "true" or null |
| negative_covid_test | string | "true" or null |
| Suspected_COVID | string | "true" or null |
| in_death_table | string | "true" or null. True if patient known to be deceased |
| age_at_visit_start_in_years_int | integer | age in years |
| length_of_stay | integer | length of stay in days |
| Race | string | e.g., "Missing/Unknown" |
| Ethnicity | string | e.g., "Missing/Unknown" |
| gender_concept_name | string | e.g., "FEMALE" |
| smoking_status | string | e.g., "Non smoker" |
| blood_type | string | e.g., "Unknown" |
| covid_status_name | string | e.g., "covid_confirmed_positive" |
| Severity_Type | string | e.g., "Mild" |
| InpatientOrED | boolean | true or false |
| Q_Score | integer | Charlson Comorbidity Index |
| BMI | double | e.g., 18.2 |
| Height | double | in meters |
| Weight | double | in kilograms |
| Testcount | integer | Number of COVID tests performed |

**Table S7.** Fields of the complete_patient_table_with_derived_scores table

##### S1.2.1 complete_patient_table_with_covariates

**Inputs:**

- `complete_patient_table_with_derived_scores` (Section S1.2)
- `covariate_list`. This is a table with true or false values (in columns) for the covariates that is derived from the condition_occurence table and the corresponding concept sets in a straightforward fashion that is omitted here for conciseness.

```
SELECT *
FROM complete_patient_table_with_derived_scores
LEFT JOIN covariate_list USING(person_id)
```

**Listing 1:** Join the covariates to the patient table.

#### S1.3 create cohort dataset

**Inputs:**

- `concept_set_members` (Sec. S1.1.3)
- `drug_era` (Sec. S1.1.1)
- `complete_patient_table_with_covariates` (Sec. S1.2.1)

The following code creates the cohort dataset we will use for statistical analysis.

```
def create_cohort_dataset( concept_set_members, drug_era,
 complete_patient_table_with_covariates) -> None:
    dat = complete_patient_table_with_covariates
    covariates_to_check = ["alcoholic_liver_damage","chronic_hepatitis","diabetes2",
    "hepatic_failure", "hypertension","ischemic_heart_disease","lupus",
    "malignant_neoplasm_lymphoid_hematopoietic_related_tissue",
    "neoplasm","nonischemic_heart_disease","vascular_dementia","alzheimers_disease",
    "cerebral_infarction","chronic_resp","dementia_associated_with_another_disease",
    "diabetes1","hepatic_fibrosis","hepatic_steatosis","hypertensive_kidney_disease",
    "nicotine_dependence","nonhypertensive_chronic_kidney_disease","other_liver_disease",
    "portal_hypertension","rheumatoid_arthritis","unspecified_dementia","psoriasis",
    "immunosuppression"]

    min_num_of_cases_to_keep_covariate = 1000 # any cov with less than 1000 True's will be
    dropped

    drug_concept_sets = ["ibuprofen-107", "oxicams-jac", "ketorolac-9", "diclofenac-28", "
   celecoxib-2", "indomethacin-72", "naproxen-4", "etodolac-36"]
    exclude_drug_concept_sets = ["aspirin-1-9", "acetaminophen-1-26"]

   # filter for COVID+
    patient_table = dat.filter(dat.positive_covid_test==True)
    drug_concept_ids = None

    for this_concept_set_name in drug_concept_sets:
        try:
            these_drug_concept_ids = get_codeset(this_concept_set_name,
            concept_set_members, get_is_most_recent=True, exclude_topicals=True,
             verbose=verbose)
        except NoConceptIdsError:
            raise Exception("I couldn't find any concept IDs for {this_concept_set_name}")

        if not drug_concept_ids:
            drug_concept_ids = these_drug_concept_ids
```

```
        else:
            drug_concept_ids = DataFrame.unionAll(drug_concept_ids, these_drug_concept_ids
    )

    # make column with on_drug
    pts_on_drug = get_pts_on_drug(patient_table, drug_era,
      drug_concept_ids, verbose=False)
    pts_on_drug = pts_on_drug.withColumnRenamed("person_id", "drug_person_id")
    patient_table = patient_table.join(pts_on_drug,
      patient_table.person_id == pts_on_drug.drug_person_id, how="leftouter")
    patient_table = patient_table.withColumn("on_drug",
      (patient_table.drug_person_id.isNotNull()))
    patient_table = patient_table.drop("drug_person_id")
    treated_pt_count = patient_table.filter(patient_table.on_drug==True).count()

    # Flag patients taking other NSAIDs (e.g. acetaminophen, aspirin)
    excluded_drug_concept_ids = None
    for this_concept_set_name in exclude_drug_concept_sets:
        # get drug concept IDs
        try:
            these_excluded_drug_concept_ids = get_codeset(this_concept_set_name,
             concept_set_members, get_is_most_recent=True, exclude_topicals=True,
              verbose=verbose)
        except NoConceptIdsError:
            raise Exception("(Excluded drug) I couldn't find any concept IDs for {
    this_concept_set_name}")
        if not excluded_drug_concept_ids:
            excluded_drug_concept_ids = these_excluded_drug_concept_ids
        else:
            excluded_drug_concept_ids = DataFrame.unionAll(excluded_drug_concept_ids,
             these_excluded_drug_concept_ids)

    pts_on_excluded_drug = get_pts_on_drug(patient_table, drug_era,
     excluded_drug_concept_ids, verbose=False)

    # get list of pts on excluded drug and not on_drug of interest
    not_on_drug_pts = patient_table.filter(patient_table.on_drug ==
      False).select("person_id")
    not_on_drug_pts = not_on_drug_pts.withColumnRenamed("person_id", "drug_person_id")

    exclude_patients = pts_on_excluded_drug.join(not_on_drug_pts, pts_on_excluded_drug.
    person_id == not_on_drug_pts.drug_person_id, how="inner").select("drug_person_id")


    # make a new column with info about on_excluded_drug, so we can analyze cohort numbers
     coherently in the next node
    patient_table = patient_table.join(exclude_patients, patient_table.person_id ==
    exclude_patients.drug_person_id, how="leftouter")
    patient_table = patient_table.withColumn("on_excluded_drug", (patient_table.
    drug_person_id.isNotNull()))
    patient_table = patient_table.drop("drug_person_id")

    # Some data cleaning on covariates
    # merge rheumatoid_arthritis_w_factor and rheumatoid_arthritis columns
    from pyspark.sql.functions import when
    patient_table = patient_table.withColumn("ra_or_ra_with_factor", when(patient_table.
    rheumatoid_arthritis_w_factor == True, True)
       .when(patient_table.rheumatoid_arthritis == True, True)
       .otherwise(False))
    patient_table = patient_table.drop("rheumatoid_arthritis", "
```

```
rheumatoid_arthritis_w_factor")
    patient_table = patient_table.withColumnRenamed("ra_or_ra_with_factor", "
rheumatoid_arthritis")

    # replace null's with False
    patient_table = patient_table.fillna(value=False,subset=covariates_to_check)

    # remove covariates with less than 1000 patients
    for cov in covariates_to_check:
        ncases = patient_table.filter(patient_table[cov] == True).count()
        if ncases < min_num_of_cases_to_keep_covariate:
            print(f"dropping {cov} column since it's less than
min_num_of_cases_to_keep_covariate ({min_num_of_cases_to_keep_covariate})")
            patient_table = patient_table.drop(cov)

    return(patient_table)
```

**Listing 2:** Create the case/control dataset

#### S1.4 drop outpatient and ed

**Inputs:**

- `create_cohort_dataset` ( S1.3; note that we omit an intermediate step that removes data from centers that do not provide BMI data)

This node performs drops outpatients as well as outpatients who were seen in an emergency department (ED) but not admitted.

In effect, this node drops visits classified as "Outpatient Visit", "Emergency Room Visit", "No matching concept", "Office Visit", "Information not available", "Telehealth", "Non-hospital institution Visit", "Observation Room", "Ambulatory Surgical Center", "Ambulatory Clinic / Center", "Laboratory Visit", "Interactive Telemedicine Service", "Ambulatory Radiology Clinic / Center", "Ambulatory Infusion Therapy Clinic / Center", "Ambulatory Dental Clinic / Center", "Ambulatory Oncology Clinic / Center", "Ambulatory Magnetic Resonance Imaging (MRI) Clinic / Center", "Ambulatory Mammography Clinic / Center", "Ambulatory Oncological Radiation Clinic / Center", "Home Visit", "Ambulatory Endoscopy Cinic / Center", "Skilled Nursing Facility".

```
drop_outpatient_and_ed <- function(create_cohort_dataset) {
    dat <- throw_out_data_partners_with_100_perc_missing_BMI
    dat <- dat[dat$visit_concept_name %in%  c("Inpatient Visit", "Inpatient Hospital",
     "Emergency Room and Inpatient Visit", "Inpatient Critical Care Facility" ),]
    return(dat)
}
```

**Listing 3:** Remove outpatients

#### S1.5 do matchit drop outpatient and ed

**Inputs:**

- `drop_outpatient_and_ed` (Section S1.4)

This node performs propensity matching on data that contains only inpatients (outpatient and outpatient emergency department have been dropped).

Todo – explain `on_excluded_drug`

```
do_matchit_drop_outpatient_and_ed <- function(drop_outpatient_and_ed) {
    covariates_to_exact_match <- NULL
    excluded_drug_col <- "on_excluded_drug"
    dat <- drop_outpatient_and_ed

    # cols we need below
    factor_cols <- c("Race", "Ethnicity", "gender_concept_name", "smoking_status",
     "InpatientOrED", "Severity_Type", "data_partner_id")
    outcomes <- c("Severity_Type", "in_death_table", "Invasive_Ventilation",
     "AKI_in_hospital", "ECMO", "InpatientOrED")
    treatment <- "on_drug"
    data_partner_col = "data_partner_id"
    patient_info_for_matching <- c("age_at_visit_start_in_years_int", "Race", "Ethnicity",
     "gender_concept_name", "smoking_status", "BMI", "Q_Score")
    comorbidities_for_matching <- c("alcoholic_liver_damage", "chronic_hepatitis",
     "diabetes2", "hepatic_failure", "hypertension", "ischemic_heart_disease",
     "lupus", "malignant_neoplasm_lymphoid_hematopoietic_related_tissue",
     "neoplasm", "nonischemic_heart_disease", "vascular_dementia",
     "alzheimers_disease", "cerebral_infarction", "chronic_resp",
      "dementia_associated_with_another_disease", "diabetes1",
      "hepatic_fibrosis", "hepatic_steatosis", "hypertensive_kidney_disease",
       "nicotine_dependence", "nonhypertensive_chronic_kidney_disease",
        "other_liver_disease", "portal_hypertension", "rheumatoid_arthritis",
         "unspecified_dementia", "psoriasis")

    # filter out patients "on_excluded_drug"
    dat <- filter(dat, dat[[excluded_drug_col]] == FALSE)

    uniq_pts = length(unique(dat$person_id))
    print(paste("found", uniq_pts, "person_ids after filtering out pts on_excluded_drug"))

    # clean/convert outcome columns into boolean
    dat$in_death_table = !is.na(dat$in_death_table) & dat$in_death_table == TRUE
    dat$Invasive_Ventilation = !is.na(dat$Invasive_Ventilation)
    dat$ECMO = !is.na(dat$ECMO)
    dat$AKI_in_hospital = !is.na(dat$AKI_in_hospital)

    cols_of_interest <- c(
        # fairly standard patient info
        "person_id", # always keep this
        outcomes,
        treatment,
        data_partner_col,
        patient_info_for_matching,
        comorbidities_for_matching)

    matchit.formula <- as.formula(paste(treatment, "~",
        paste(c(patient_info_for_matching, comorbidities_for_matching, data_partner_col),
         collapse="+")))
    matched.data <- do_matchit(data=dat, cols_of_interest=cols_of_interest,
     matchit.formula=matchit.formula, factor_cols=factor_cols,
     matchit.method="nearest", verbose=verbose,
     exact_matching=covariates_to_exact_match, do.love=TRUE, make_table_one=FALSE,
      vim.margin.plot.cols=c("age_at_visit_start_in_years_int", "BMI"))

    if (!exists("matched.data") || is.null(matched.data)){
        stop("matched.data is null after matching")
    }

    return(matched.data)
```

```
}
```

**Listing 4:** propensity matching

#### S1.6 glm regression inpatient no ed mild mild ed moderate versus severe dead

**Inputs:**

- `do_matchit_drop_outpatient_and_ed` (Section S1.5)

The following code expects as input a data table with matched patients and controls. It uses a convenience function called `set_cols_as_factors` that converts columns to factor using the following command for each of the selected columns.

`dat[[x]] <- as.factor(dat[[x]])`

```
glm_regression_inpatient_no_ed_mild_mild_ed_moderate_versus_severe_dead <- function(do_matchit_drop_outpatient_and_ed) {
    matched.data <- do_matchit_drop_outpatient_and_ed
    factor_cols <- c("Race", "Ethnicity", "gender_concept_name", "smoking_status",
     "InpatientOrED", "Severity_Type", "data_partner_id")
    glm_outcomes <- c("Severity_Type", "in_death_table", "Invasive_Ventilation",
     "AKI_in_hospital", "ECMO")

    matched.data <- set_cols_as_factors(matched.data, factor_cols, verbose=FALSE)

    matched.data[["Severity_Type"]] <- as.numeric(matched.data[["Severity_Type"]] %in% c("Severe", "Dead_w_COVID"))

    # these were selected by the code in "select_covariates" node
    covariates_always_keep <- c("on_drug", "age_at_visit_start_in_years_int", "Race",
     "Ethnicity", "gender_concept_name", "smoking_status", "BMI")
    selected_covariates <- c("alcoholic_liver_damage",
                             "alzheimers_disease",
                             "cerebral_infarction",
                             "chronic_resp",
                             "diabetes1",
                             "diabetes2",
                             "hepatic_failure",
                             "hepatic_fibrosis",
                             "hypertensive_kidney_disease",
                             "ischemic_heart_disease",
                             "lupus",
                             "malignant_neoplasm_lymphoid_hematopoietic_related_tissue",
                             "neoplasm",
                             "nicotine_dependence",
                             "nonhypertensive_chronic_kidney_disease",
                             "nonischemic_heart_disease",
                             "other_liver_disease",
                             "portal_hypertension",
                             "psoriasis",
                             "Q_Score",
                             "unspecified_dementia",
                             "vascular_dementia")

    covariate_terms_for_formula <- paste(c(covariates_always_keep, selected_covariates), collapse="+")
```

```
    # do logistic regression
    all_glms <- lapply(glm_outcomes, function(outcome){
        logit.formula <- as.formula(paste(outcome, "~", covariate_terms_for_formula))
        logit <- NULL
        try(logit <- do_glm(dat=matched.data, glm.formula=logit.formula))
        return(logit)
    })
    print(all_glms)

    # make table with summary
    parsed_data <- lapply(all_glms, function(logit){
        logit_summary <- summary(logit)
        logit_coef <- coef(logit_summary)
        all_coeff_data <- cbind(logit_coef, data.frame("OR"=exp(logit_coef[,'Estimate'])),
         exp(confint(logit)))
        all_coeff_data <- all_coeff_data["on_drugTRUE",] %>%
              dplyr::select(-c("Estimate", "Std. Error", "z value"))
        return(all_coeff_data)
    })
    print(cbind(glm_outcomes, do.call(rbind, parsed_data)))

    return(NULL)
}
```

**Listing 5:** glm regression.

#### S1.7 generate outcome counts table inpatients no ed

**Inputs:**

- `do_matchit_drop_outpatient_and_ed` (Section S1.5)

This script is used to generate output as such as the following.

```
# A tibble: 3 x 6
# Groups:   on_drug [1]
  on_drug Severity_Type      n  perc    n1 perc1
  <lgl>   <ord>          <int> <dbl> <int> <dbl>
1 TRUE    Moderate       18023 91.3  16972 86.0
2 TRUE    Severe           776  3.93  1047  5.30
3 TRUE    Dead_w_COVID     947  4.80  1727  8.75
# A tibble: 4 x 5
  outcome                    n  perc    n1  perc1
  <chr>                  <int> <dbl> <int>  <dbl>
1 in_death_table           947 4.80   1727   8.75
2 Invasive_Ventilation    1150 5.82   1867   9.46
3 AKI_in_hospital         1729 8.76   2437  12.3
4 ECMO                      54 0.273   109   0.552
```

```
generate_outcome_counts_table_inpatients_no_ed <-
 function(do_matchit_drop_outpatient_and_ed) {
    matched.data <- do_matchit_drop_outpatient_and_ed
    secondary_outcomes <- c("Severity_Type", "in_death_table", "Invasive_Ventilation",
     "AKI_in_hospital", "ECMO")
    factor_cols <- c("Race", "Ethnicity", "gender_concept_name", "smoking_status",
```

```
    "InpatientOrED", "Severity_Type", "data_partner_id")
    matched.data <- set_cols_as_factors(matched.data, factor_cols, verbose=FALSE)
    matched.data <- fix_ordinal_outcome(matched.data)

    summaries <- lapply(secondary_outcomes, function(this.outcome){
        this.outcome.tab <- matched.data %>% dplyr::group_by(on_drug) %>% dplyr::count(!!
    sym(this.outcome))
        this.outcome.tab <- this.outcome.tab %>% dplyr::mutate(perc = 100 * n / sum(n))
        this.outcome.tab <- cbind(this.outcome.tab %>% filter(on_drug), this.outcome.tab
    %>% filter(!on_drug))
        return(this.outcome.tab)
    })

    # severity type data:
    std <- summaries[[1]] %>% dplyr::select(Severity_Type, n, perc, n1, perc1)
    print(std)

    secondary_outcomes <- lapply(summaries[2:5], function(dat){
        # getting rid of the "FALSE" row because it's unnecessary, and using the name of
    the 2nd column as the outcome
        dat <- dat[dat[,2]==TRUE,] %>% mutate(outcome=colnames(dat)[2]) %>% dplyr::select(
    outcome, n, perc, n1, perc1) %>% ungroup() %>% dplyr::select(-on_drug)
        return(dat)
    })
    print(dplyr::bind_rows(secondary_outcomes))
    return(NULL)
}
```

**Listing 6:** Generate outcome counts.

### References


[1] Laila Carolina Abu Esba, Rahaf Ali Alqahtani, Abin Thomas, Nour Shamas, Lolowa Alswaidan, and Gahdah Mardawi. Ibuprofen and nsaid use in covid-19 infected patients is not associated with worse outcomes: A prospective cohort study. *Infectious diseases and therapy*, 10:253–268, March 2021.

[2] Nasser Malekpour Alamdari, Siamak Afaghi, Fatemeh Sadat Rahimi, Farzad Esmaeili Tarki, Sasan Tavana, Alireza Zali, Mohammad Fathi, Sara Besharat, Leyla Bagheri, Fatemeh Pourmotahari, Seyed Sina Naghibi Irvani, Ali Dabbagh, and Seyed Ali Mousavi. Mortality risk factors among hospitalized covid-19 patients in a major referral center in iran. *The Tohoku journal of experimental medicine*, 252:73–84, September 2020.

[3] Eilidh Bruce, Fenella Barlow-Pay, Roxanna Short, Arturo Vilches-Moraga, Angeline Price, Aine McGovern, Philip Braude, Michael J. Stechman, Susan Moug, Kathryn McCarthy, Jonathan Hewitt, Ben Carter, and Phyo Kyaw Myint. Prior routine use of non-steroidal anti-inflammatory drugs (nsaids) and important outcomes in hospitalised patients with covid-19. *Journal of clinical medicine*, 9, August 2020.

[4] Joht Singh Chandan, Dawit Tefra Zemedikun, Rasiah Thayakaran, Nathan Byne, Samir Dhalla, Dionisio Acosta-Mena, Krishna M. Gokhale, Tom Thomas, Christopher Sainsbury, Anuradhaa Subramanian, Jennifer Cooper, Astha Anand, Kelvin O. Okoth, Jingya Wang, Nicola J. Adderley, Thomas Taverner, Alastair K. Denniston, Janet Lord, G. Neil Thomas, Christopher D. Buckley, Karim Raza, Neeraj Bhala, Krishnarajah Nirantharakumar, and Shamil Haroon. Nonsteroidal antiinflammatory drugs and susceptibility to covid-19. *Arthritis & Rheumatology (Hoboken, N.J.)*, 73:731–739, May 2021.

[5] Min Hyuk Choi, Hyunmin Ahn, Han Seok Ryu, Byung-Jun Kim, Joonyong Jang, Moonki Jung, Jinuoung Kim, and Seok Hoon Jeong. Clinical characteristics and disease progression in early-stage covid-19 patients in south korea. *Journal of clinical medicine*, 9, June 2020.

[6] Thomas M. Drake, Cameron J. Fairfield, Riinu Pius, Stephen R. Knight, Lisa Norman, Michelle Girvan, Hayley E. Hardwick, Annemarie B. Docherty, Ryan S. Thwaites, Peter J. M. Openshaw, J. Kenneth Baillie, Ewen M. Harrison, Malcolm G. Semple, and ISARIC4C. Investigators. Nonsteroidal anti-inflammatory drug use and outcomes of covid-19 in the isaric clinical characterisation protocol uk cohort: a matched, prospective cohort study. *The Lancet. Rheumatology*, 3:e498–e506, July 2021.

[7] Milena Gianfrancesco, Kimme L. Hyrich, Sarah Al-Adely, Loreto Carmona, Maria I. Danila, Laure Gossec, Zara Izadi, Lindsay Jacobsohn, Patricia Katz, Saskia Lawson-Tovey, Elsa F. Mateus, Stephanie Rush, Gabriela Schmajuk, Julia Simard, Anja Strangfeld, Laura Trupin, Katherine D. Wysham, Suleman Bhana, Wendy Costello, Rebecca Grainger, Jonathan S. Hausmann, Jean W. Liew, Emily Sirotich, Paul Sufka, Zachary S. Wallace, Jinoos Yazdany, Pedro M. Machado, Philip C. Robinson, and C. O. V. I. D.-19 Global Rheumatology Alliance. Characteristics associated with hospitalisation for covid-19 in people with rheumatic disease: data from the covid-19 global rheumatology alliance physician-reported registry. *Annals of the rheumatic diseases*, 79:859–866, July 2020.

[8] Shruti Gupta, Salim S. Hayek, Wei Wang, Lili Chan, Kusum S. Mathews, Michal L. Melamed, Samantha K. Brenner, Amanda Leonberg-Yoo, Edward J. Schenck, Jared Radbel, Jochen Reiser, Anip Bansal, Anand Srivastava, Yan Zhou, Anne Sutherland, Adam Green, Alexandre M. Shehata, Nitender Goyal, Anitha Vijayan, Juan Carlos Q. Velez, Shahzad Shaefi, Chirag R. Parikh, Justin Arunthamakun, Ambarish M. Athavale, Allon N. Friedman, Samuel A. P. Short, Zoe A. Kibbelaar, Samah Abu Omar, Andrew J. Admon, John P. Donnelly, Hayley B. Gershengorn, Miguel A. Hernán, Matthew W. Semler, David E. Leaf, and S. T. O. P.-C. O. V. I. D. Investigators. Factors associated

with death in critically ill patients with coronavirus disease 2019 in the us. *JAMA internal medicine*, 180:1436–1447, November 2020.

[9] Jong-Moon Hwang, Ju-Hyun Kim, Jin-Sung Park, Min Cheol Chang, and Donghwi Park. Neurological diseases as mortality predictive factors for patients with covid-19: a retrospective cohort study. *Neurological sciences : official journal of the Italian Neurological Society and of the Italian Society of Clinical Neurophysiology*, 41:2317–2324, September 2020.

[10] Z. Imam, F. Odish, I. Gill, D. O'Connor, J. Armstrong, A. Vanood, O. Ibironke, A. Hanna, A. Ranski, and A. Halalau. Older age and comorbidity are independent mortality predictors in a large cohort of 1305 covid-19 patients in michigan, united states. *Journal of internal medicine*, 288:469–476, October 2020.

[11] Kristian Kragholm, Thomas A. Gerds, Emil Fosbøl, Mikkel Porsborg Andersen, Matthew Phelps, Jawad H. Butt, Lauge Østergaard, Casper N. Bang, Jannik Pallisgaard, Gunnar Gislason, Morten Schou, Lars Køber, and Christian Torp-Pedersen. Association between prescribed ibuprofen and severe covid-19 infection: A nationwide register-based cohort study. *Clinical and translational science*, 13:1103–1107, November 2020.

[12] Lars Christian Lund, Kasper Bruun Kristensen, Mette Reilev, Steffen Christensen, Reimar Wernich Thomsen, Christian Fynbo Christiansen, Henrik Støvring, Nanna Borup Johansen, Nikolai Constantin Brun, Jesper Hallas, and Anton Pottegård. Adverse outcomes and mortality in users of non-steroidal anti-inflammatory drugs who tested positive for sars-cov-2: A danish nationwide cohort study. *PLoS medicine*, 17:e1003308, September 2020.

[13] Jungchan Park, Seung-Hwa Lee, Seng Chan You, Jinseob Kim, and Kwangmo Yang. Non-steroidal anti-inflammatory agent use may not be associated with mortality of coronavirus disease 19. *Scientific reports*, 11:5087, March 2021.

[14] E. Rinott, E. Kozer, Y. Shapira, A. Bar-Haim, and I. Youngster. Ibuprofen use and clinical outcomes in covid-19 patients. *Clinical microbiology and infection : the official publication of the European Society of Clinical Microbiology and Infectious Diseases*, 26:1259.e5–1259.e7, September 2020.

[15] Aditya Sahai, Rohan Bhandari, Matthew Godwin, Thomas McIntyre, Mina K. Chung, Jean-Pierre Iskandar, Hayaan Kamran, Essa Hariri, Anu Aggarwal, Robert Burton, Ankur Kalra, John R. Bartholomew, Keith R. McCrae, Ayman Elbadawi, James Bena, Lars G. Svensson, Samir Kapadia, and Scott J. Cameron. Effect of aspirin on short-term outcomes in hospitalized patients with covid-19. *Vascular medicine (London, England)*, 26:626–632, December 2021.

[16] Angel Ys Wong, Brian MacKenna, Caroline E. Morton, Anna Schultze, Alex J. Walker, Krishnan Bhaskaran, Jeremy P. Brown, Christopher T. Rentsch, Elizabeth Williamson, Henry Drysdale, Richard Croker, Seb Bacon, William Hulme, Chris Bates, Helen J. Curtis, Amir Mehrkar, David Evans, Peter Inglesby, Jonathan Cockburn, Helen I. McDonald, Laurie Tomlinson, Rohini Mathur, Kevin Wing, Harriet Forbes, Rosalind M. Eggo, John Parry, Frank Hester, Sam Harper, Stephen Jw Evans, Liam Smeeth, Ian J. Douglas, Ben Goldacre, and OpenSA. F. E. L. Y. Collaborative. Use of non-steroidal anti-inflammatory drugs and risk of death from covid-19: an opensafely cohort analysis based on two cohorts. *Annals of the rheumatic diseases*, 80:943–951, July 2021.

[17] Melissa A Haendel, Christopher G Chute, Tellen D Bennett, David A Eichmann, Justin Guinney, Warren A Kibbe, Philip R O Payne, Emily R Pfaff, Peter N Robinson, Joel H Saltz, Heidi Spratt, Christine Suver, John Wilbanks, Adam B Wilcox, Andrew E Williams, Chunlei Wu, Clair Blacketer, Robert L Bradford, James J Cimino, Marshall Clark, Evan W Colmenares, Patricia A Francis, Davera Gabriel, Alexis Graves, Raju Hemadri, Stephanie S Hong, George Hripscak, Dazhi Jiao, Jeffrey G

Klann, Kristin Kostka, Adam M Lee, Harold P Lehmann, Lora Lingrey, Robert T Miller, Michele Morris, Shawn N Murphy, Karthik Natarajan, Matvey B Palchuk, Usman Sheikh, Harold Solbrig, Shyam Visweswaran, Anita Walden, Kellie M Walters, Griffin M Weber, Xiaohan Tanner Zhang, Richard L Zhu, Benjamin Amor, Andrew T Girvin, Amin Manna, Nabeel Qureshi, Michael G Kurilla, Sam G Michael, Lili M Portilla, Joni L Rutter, Christopher P Austin, Ken R Gersing, and N3C Consortium. The national covid cohort collaborative (n3c): Rationale, design, infrastructure, and deployment. *Journal of the American Medical Informatics Association : JAMIA*, 28:427–443, March 2021.

[18] Tellen D Bennett, Richard A Moffitt, Janos G Hajagos, Benjamin Amor, Adit Anand, Mark M Bissell, Katie Rebecca Bradwell, Carolyn Bremer, James Brian Byrd, Alina Denham, Peter E DeWitt, Davera Gabriel, Brian T Garibaldi, Andrew T Girvin, Justin Guinney, Elaine L Hill, Stephanie S Hong, Hunter Jimenez, Ramakanth Kavuluru, Kristin Kostka, Harold P Lehmann, Eli Levitt, Sandeep K Mallipattu, Amin Manna, Julie A McMurry, Michele Morris, John Muschelli, Andrew J Neumann, Matvey B Palchuk, Emily R Pfaff, Zhenglong Qian, Nabeel Qureshi, Seth Russell, Heidi Spratt, Anita Walden, Andrew E Williams, Jacob T Wooldridge, Yun Jae Yoo, Xiaohan Tanner Zhang, Richard L Zhu, Christopher P Austin, Joel H Saltz, Ken R Gersing, Melissa A Haendel, and Christopher G Chute. The national covid cohort collaborative: Clinical characterization and early severity prediction. *medRxiv : the preprint server for health sciences*, January 2021.

[19] George Hripcsak, Jon D. Duke, Nigam H. Shah, Christian G. Reich, Vojtech Huser, Martijn J. Schuemie, Marc A. Suchard, Rae Woong Park, Ian Chi Kei Wong, Peter R. Rijnbeek, Johan van der Lei, Nicole Pratt, G. Niklas Norén, Yu-Chuan Li, Paul E. Stang, David Madigan, and Patrick B. Ryan. Observational health data sciences and informatics (ohdsi): Opportunities for observational researchers. *Studies in health technology and informatics*, 216:574–578, 2015.
